# “Us with them”: Co-designing a caesarean section consent and debriefing intervention in West Cameroon Co-design for better consent and debriefing for caesarean section

**DOI:** 10.64898/2026.06.17.26355846

**Authors:** Jovanny Tsuala Fouogue, Miho Sato, Louise Tina Day, Mitsuaki Matsui, William Carter Djuatio Kenne, Bruno kenfack, Lenka Beňová, Veronique Filippi

**Author notes:** Corresponding author (JTF).

## Abstract

**Background:** Women-centred maternity care is a rights issue that determines the use of services. Such care ensures responsiveness to women’s needs which is enacted through shared decision-making, review and response. In the West Region of Cameroon, informed consent (IC) and Debriefing for caesarean section (c-section) have been shown to be suboptimal or absent. This paper describes the participatory design of a quality-improvement hospital-based intervention.

**Methods:** From February to May 2025, we conducted a co-design process with three groups of stakeholders: 59 post c-section women and community representatives, 78 frontline c-section providers, and 29 directors of public and private hospitals. We followed four phases: planning, conducting, evaluating, and reporting. The conduct phase comprised five all-day workshops with post c-section women and community representatives, followed by five all-day workshops with the c-section providers. Finally, we held an 11^th^ workshop with the hospital directors to scrutinize suggested interventions, evaluate their feasibility, and establish a consensus on their components. We described the intervention using the TIDieR (Template for Intervention Description and Replication) checklist. We documented the co-design process, using open-ended narratives to delineate interventions, and carried out real-time synthesis on visual aids (whiteboards and flipcharts). Intervention feasibility was quantified using a structured *ad hoc* matrix, while insights on facilitators and barriers were captured through qualitative free-text entries. We coupled data collection with constant comparison and triangulation through contemporaneous field notes, photographic documentation, and thematic mapping of stakeholders’ perceptions and interactive dynamics.

**Results:** Participants’ perspectives on the co-design were positive, and their motivation were very high although less than 50% reported previous involvement in co-design processes. More than 80% of participants found rated the co-design process as either good or very good. The final intervention comprised four components: (i) an in-service training; (ii) a standard operating procedure including a harmonised consent form and debriefing checklist; (ii) systematic supportive supervision, monitoring & evaluation; and (iv) a routine clinical audit. Each group of stakeholders upheld specific dimensions of the consent and debrief intervention. Post c-section women and community members emphasized emotional support, written discharge advice after debriefing, and zero tolerance of suboptimal consent and debriefing practices. Frontline c-section providers insisted on robust documentation for medico-legal protection. Hospitals Directors emphasized capacity-building and cultural friendliness. All the groups supported woman’s autonomous decision making. The intervention feasibility was rated high or very high by hospital directors except for the financial, infrastructural and technical domains.

**Conclusion:** This co-design process yielded a context-specific, multi-component intervention that was well accepted and deemed feasible across stakeholders. It provides a methodological approach to strengthening informed consent and debriefing as core elements of women-centred, accountable maternity care, and warrants implementation.

## Introduction

Caesarean section (C-section) delivers the greatest benefit when high-quality technical care is embedded in respectful, dignified and person-centred care (1). Recent increases in country prevalences, frequently exceeding optimal levels, have been linked to the use of non-medical indications and suggest a pressing need for robust ethical safeguards(1–5). The issue also prevails in low-prevalence settings where health systems develop pockets where non-medically indicated c-sections are practiced (6,7). A decade ago, the World Health Organization (WHO) framework for quality of maternal and newborn care introduced women-centred dimensions alongside technical aspects of clinical care(8). To support countries in operationalising this paradigm shift toward people-centred and integrated care in the emerging era of universal health coverage (UHC), the WHO has developed a strategy based on five interdependent pillars centred on co-production (9). Regardless of context, a core principle of people-centred care is that services must be ethical, including respect for individuals’ right to make autonomous and informed decisions about their health care. With respect to c-section (and maternal health more broadly) community involvement in co-production of health care can be enacted through the participatory design of clinical guidelines on communication with women and their next-of-kin before and after the procedure.

In clinical practice, informed consent (IC) is a medico-legal requirement prior to any medical, obstetrical, or surgical procedure; it entails more than simply obtaining a signature and includes the honest disclosure of understandable information to a patient whose decision-making capacity has been confirmed, sufficient time for reflection, answers to any questions, and a clear request for authorization to carry out the procedure (10,11). Debriefing is a structured two-way communication session that provides an account of the procedure, assesses the patient’s experience, discusses its implications for future health, and outlines a follow-up plan (12). Emerging literature on IC and Debriefing for c-section in sub-Saharan Africa indicates low prevalence and women’s dissatisfaction with communication (4,13–23). Indeed, women complain of receiving little or no information before and after c-section. Likewise, published guidelines related to IC and Debriefing for c-section in sub-Saharan African are also scarce (12).

Cameroon is a lower-middle income country in sub-Saharan Africa country with a national c-section proportion of 3.5% (24,25). The low technical performance of the procedure, combined with too often disrespectful, undignified, culturally insensitive, content-poor practices of consent and debriefing, fuels mistrust among women and communities toward hospitals, particularly public ones. Moreover, there are widespread violations of patients’ (including women) rights in health facilities across the country and frequent controversies related to poor quality of c-section service delivery (26–30). These issues also prevail in the West Region which is one of the ten administrative regions with one the highest c-section proportions (6.2% - 10.0%) (24,31). Mindful of these circumstances and knowing that any responsive strategy to improve the quality of c-section should involve women, the most senior public health official in the Region (Regional Delegate) supported the co-design of a hospital-based intervention to improve IC and debriefing for c-section.

Participatory approaches in the design, planning, and delivery of public health interventions align with people-centred care and include co-design, co-production, and co-creation of interventions (32). The defining feature of these three approaches is the participation of communities as full actors who contribute their experiential knowledge to the cascade of public health intervention to enhance the relevance, acceptability, and impact of health interventions. Important distinctions are often made between these three concepts or approaches. Co-creation refers to collaboratively defining/identifying and developing responses to a health problem, whereas co-design involves collaboratively defining the solutions to a predefined health problem; and co-production refers to the collaborative implementation of a predefined solution (33). In Cameroon, the scientific literature on participatory development of clinical practice guidelines in reproductive health is limited (33–37).

To improve the quality of IC and Debriefing for c-section in the West Region of Cameroon, drawing upon the results of our formative research on consent and debriefing for c-section, we co-designed a context-sensitive hospital-based intervention currently under consideration for implementation. This paper we describe the processes, stakeholders’ feedback and the final intervention.

## Methods

### Participants’ selection and roles

We employed a systematic purposive sampling strategy to select stakeholders from the 1^st^ March 2025 to 30^th^ May 2025, across three distinct categories within the twenty health districts of the West Region. These included: prospective clinical end-users (frontline c-section providers), intended beneficiaries (women with a history of c-section and women’s groups leaders), and hospital leadership (directors). Participants were selected based on their involvement in our preliminary exploration of the routine practices, lived experiences, and expectations towards consent and debriefing for c-section in the Region (38–40). In addition, all the co-authors were involved in the conceptualisation of the co-design process and two of them (JTF and WCDK) took part in all the sessions. Stakeholders’ specific roles in the co-design process included: (i) researchers summarising the findings of the formative research and chairing/facilitating the meetings, (ii) intended beneficiaries and (iii) prospective end-users elaborating proposed IC and debriefing interventions; and (iv) hospitals directors assessing the feasibility of the proposed interventions. The meetings with intended beneficiaries were chaired by one researcher (JTF) and facilitated by another one (WCDK). The feasibility appraisal seminar was chaired by the Regional Delegate for public health and facilitated by the regional sexual and reproductive health lead officer the regional communication lead officer and the lead researcher (JTF).

### Period and sites

We conducted the co-design process from February to May 2025. Deliberations with intended beneficiaries and prospective end-users were divided into five geographical clusters: Mbouda (serving Mbouda, Batcham, Galim, Penka-Michel, and Dschang health districts), Foumban (serving Foumban, Malantouen, Bangourain health districts), Foumbot (serving Foumbot, Kouoptamo and Massangam health districts), Bandjoun (Bandjoun, Bamendjou, Baham, and Mifi health districts), and Bafang (serving Bafang, Bangangte, Bandja, Kekem, and Santchou health districts). It is worth noting that, due to the shortage obstetricians-gynaecologists (only a dozen in 2025) in the Region, most of the roughly 6000 yearly c-section procedures were done by non-specialist physicians and non-physician clinicians while IC was routinely sought by midwives and nurses. The feasibility appraisal seminar was held in Bafoussam, the regional capital, and brought together hospital directors to validate the proposed interventions. Supplement 1 shows these cities on a map of the West Region.

### Theoretical frameworks

During the formulation phase, the strategic rationale for each intervention was aligned with the five pillars of the WHO Framework on Integrated People-Centred Health Services: empowering individuals and communities, strengthening accountability, reorienting the model of care, coordinating multisectoral services, and fostering an enabling environment. Concurrently, the proposed interventions were mapped onto the WHO Health Systems Building Blocks (governance, financing, workforce, information systems, access to medicines/technologies and service delivery) (41). This approach ensured that the intervention addressed relevant health system components while remaining consistent with women’s needs and preferences. Additional participatory / co-design frameworks were consulted where relevant to inform specific aspects of the design, planning, conduct and evaluation. To support clear reporting and facilitate replication, the final intervention was described using the TIDieR (Template for Intervention Description and Replication) checklist (42). The TIDIeR is a guide and reporting checklist that was developed to improve the completeness of reporting, and ultimately the replicability, of intervention.

### Procedure

#### A three-phase cascade of workshops

The study employed a phased methodological approach, comprising a two-step intervention formulation phase, which was followed by a single-step operational feasibility appraisal and validation (Figure 2). The first phase comprised five intensive, all-day co-design workshops with intended beneficiaries, followed by a second series of five all-day workshops with prospective clinical end-users. This process culminated in a regional feasibility assessment and validation seminar with hospital directors to scrutinize findings, establish a consensus on the final interventions. At the start of each workshop, we presented a literature review of evidence-based interventions and the results of a multi-centre formative assessment of c-section consent and debriefing practices across 20 hospitals in the Region. To ensure evidence-informed deliberations, empirical findings from these preliminary phases were systematically presented to stakeholders at the outset of each session to contextualize the co-design activities as well as being briefed on the priorities established by beneficiaries in the previous phase.

**Figure 1.**
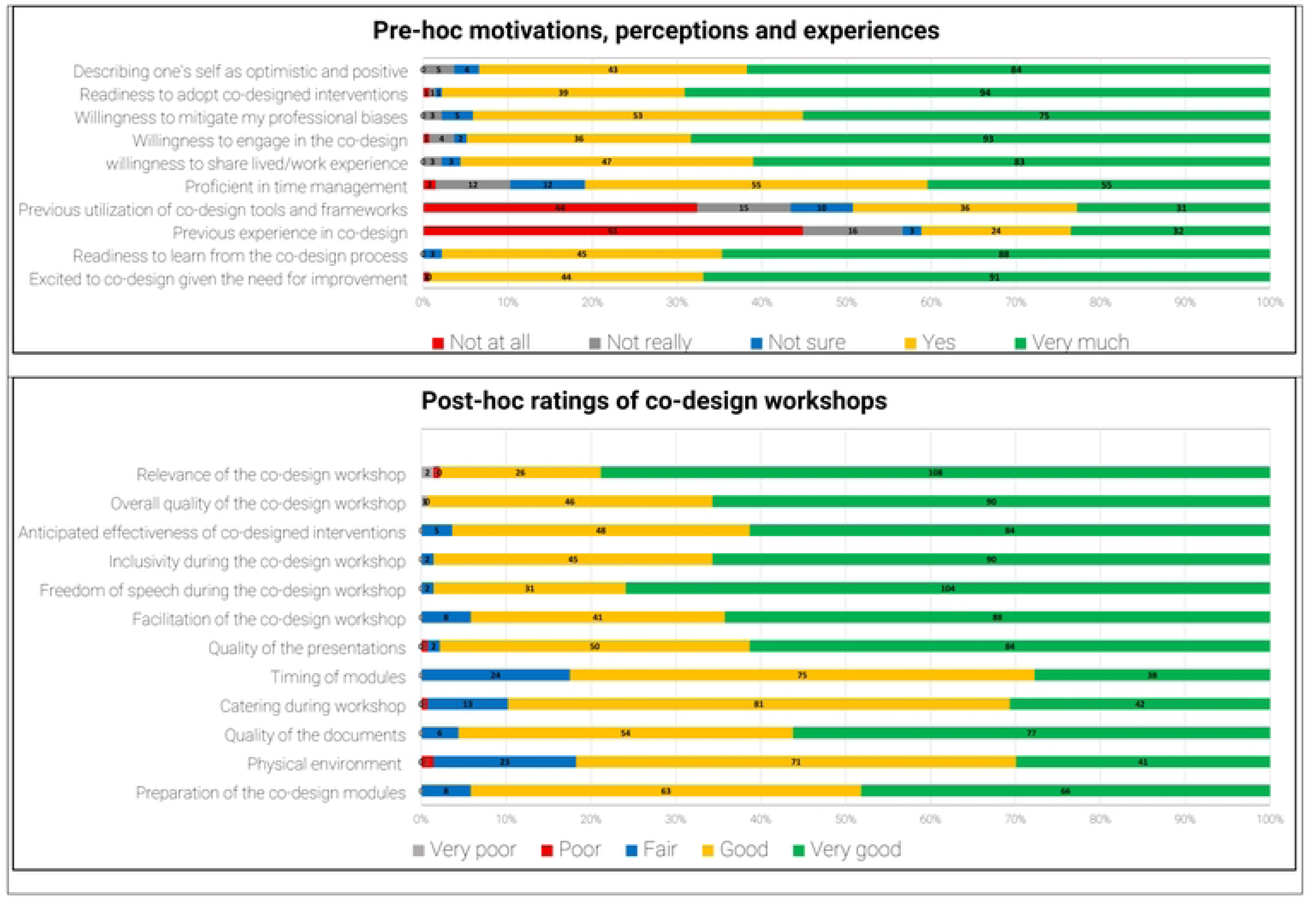
Participants’’ pre-hoc opinions and post-hoc assessment of the co-design workshops for an intervention to improve informed consent and debriefing for caesarean section: in West Cameroon. N = 137.

**Figure 2.**
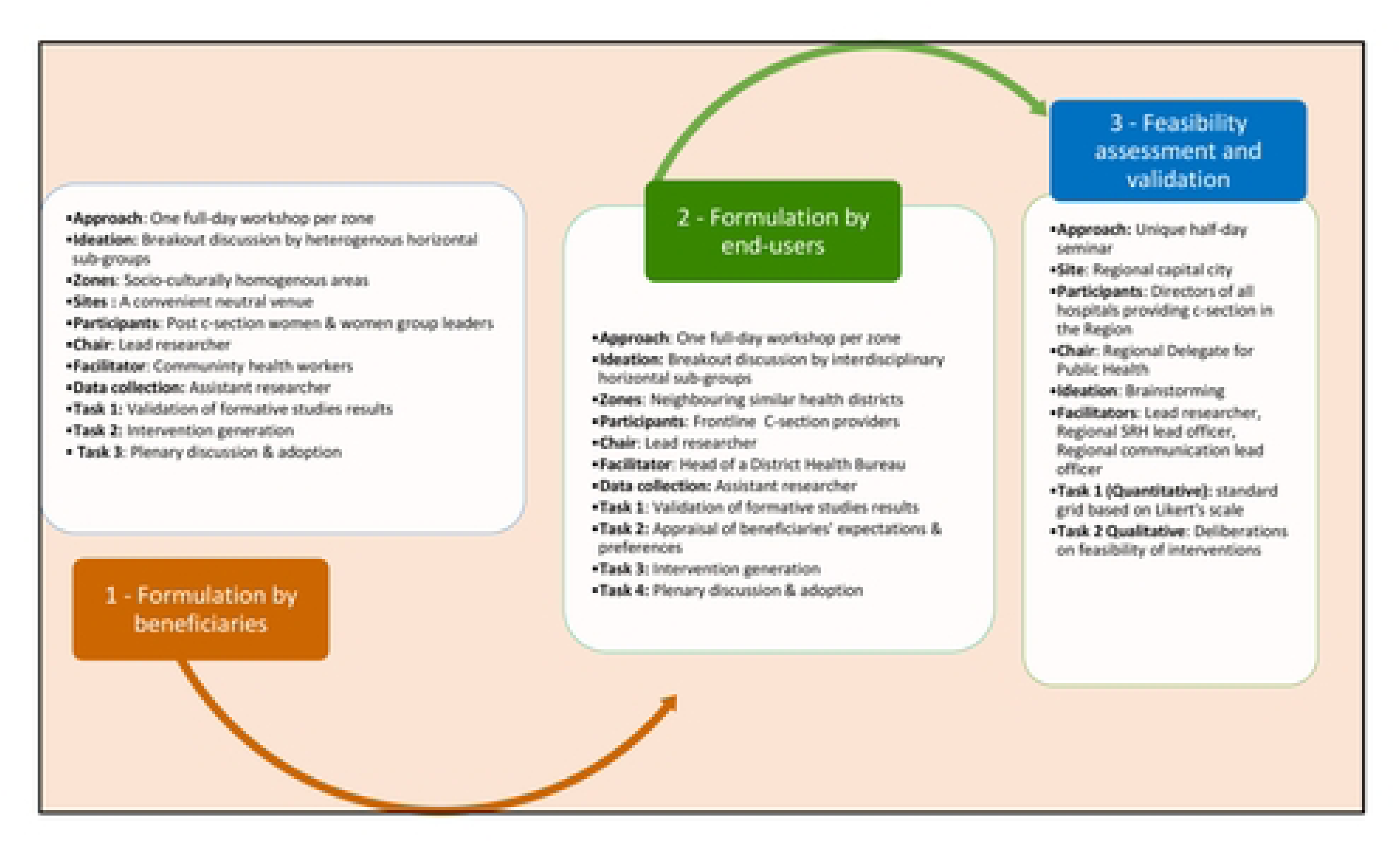
Overview of the co-design processes of a co-design of a hospital-based complex intervention to improve the quality of informed consent and debriefing for caesarean section

During the first and second phases, co-designers critically reviewed and validated the synthesized formative findings. They subsequently convened in heterogeneous breakout discussion groups (example: a male anaesthetist nurse, a labour room midwife, a female scrub nurse, and a male general practitioner from different hospitals and age groups) to formulate solutions aimed at aligning c-section consent and debriefing processes with women’s needs and preferences. These proposed solutions, alongside their strategic rationales, were refined through iterative plenary deliberations to delineate their objectives, formats, implementers, targets, contents, indicators and mechanisms. The resulting intervention components were then codified according to TIDieR checklist in view of the final appraisal by hospital directors.

In the final phase, the hospital directors conducted a feasibility appraisal of the proposed interventions preceded by a briefing on the preliminary formative findings and the co-designed prototypes which were thoroughly discussed in plenary.

#### Facilitation, group dynamics and conflict management

Facilitation was jointly done by a social scientist and an Obstetrician-Gynaecologist. The fundamental principles of participative approaches were presented and agreed upon at the beginning of every meeting. Creative thinking was fostered through the use of diverse breakout discussion groups, with reporting responsibilities intentionally assigned to less-vocal participants to mitigate the dominance bias (43–45). The outputs of breakout groups were subsequently synthesized through plenary deliberations to achieve consensual adoption through a simple majority vote provided there was no blocking from the minority. Where this consensus was not possible on a solution item, at least one different version was adopted to address the underpinnings of the disagreement. Most often, these underpinnings were specific to the context: facility management, difference between providers specialties, and social norms. Facilitation of the feasibility assessment seminar was reinforced by the participation of the communication and sexual and reproductive health officers at the Regional Delegation for Public Health, leveraging their expertise in executive-level mediation.

We held separate sessions for each group of stakeholders (prospective clinical end-users and intended beneficiaries) to minimise the effects of structural power asymmetries. We further stratified workshops with community stakeholders by gender. Consistent with the co-design principles, as facilitators, we established an enabling atmosphere that encourage constructive dissent and collaborative problem-solving.

### Data collection

JTF and WCDK collected data to: (i) capture participants’ characteristics, (ii) document the processes during the sessions, (iii) capture participants’ motivations, perceptions and experiences regarding the co-design before the workshops and their ratings thereafter, (iv) describe the co-designed interventions, (v) capture the feasibility assessment and (vi) compile the reported anticipated implementation facilitators and barriers. For each of these goals, we used a dedicated tool and approach. We collected participants characteristics with a standardized paper self-administered form immediately after the signature of the informed consent at the beginning of each session. (63) We captured participants pre- and post-workshop views through an anonymous paper semi-structured questionnaire (Supplement 2) inspired by the New South Wales Government Agency for Clinical Innovation co-design toolkit and adapted to the context (46,47). We documented the processes and interactive dynamics by real-time synthesis on visual aids (whiteboards and flipcharts), field notes, photographic documentation, and thematic mapping using sticky notes.

We collated the co-designed interventions under the form of free texts purposefully written to fit the sections of the TIDieR template (42). For the feasibility assessment we used a semi-structured form where participants followed a Likert’s scale to rate eight domains inspired by James Hall’s foundational TELOS (Technological, Economic, Legal, Organizational, Scheduling) framework (perceived relevance, priority, financial viability, technical robustness, operational logistics, implementation timelines, policy alignment, and socio-cultural acceptability) and used free text to delineate barriers and facilitators following a SWOT (Strengths, Weaknesses, Opportunities and Threats) framing (48,49).

### Data processing and analysis

We entered, checked and cleaned the participants’ socio-demographic characteristics and their feasibility ratings into an Excel sheet (Microsoft office®). We scanned and uploaded the semi-structured questionnaires containing their reported high and lows of the proceedings as well facilitators and barriers in portable document formats into a secured server. We did the same for the written descriptions of interventions crafted during breakout group work.

Using a framework-oriented approach, we synthesized data from the initial co-design steps, mapping stakeholders’ proposals onto the WHO Health Systems Building Blocks and Integrated People-Centred Health Services frameworks,

We use constant comparison to triangulate between our field notes, and participants’ answers regarding facilitators and barriers, high and low points of the proceedings and feasibility ratings. This permitted to delineate the core underpinnings of the interventions, while concurrently elucidating context-specific similarities and divergences. Data from the feasibility assessment were analysed through a parallel mixed-methods approach: quantitative appraisals were summarized via median Likert’s scores, while qualitative insights were systematically categorized using the PROSECO framework. The PROSECO (PROcesS Evaluation framework for CO-creation) framework is a systematic approach designed to assess the success of co-creation processes in the field of public health interventions. It comprises 37 items grouped in 5 core dimensions namely: delivery, participation, experience, context and impact (46).

### Ethical considerations

Ethical approval was granted by the Institutional Review Board of the London School of Hygiene & Tropical Medicine (Ref: 29898) and the Regional Ethics Committee for Human Health Research of the West Region, Cameroon (Ref: No/984/25/10/2023/CE/CRERSH-OU/VP). All participants provided written informed consent prior to enrolment. All data were labelled by the names of corresponding districts and participants categories rather than participants’ names.

## Results

### Co-designers’ characteristics

Participants’ characteristics are depicted in Table 1. Most of the c-section frontline providers (66.7%) and community members (90.0%) were women while men made the bulk of hospitals directors (79.3%). Physicians made up 10.3% of the c-section frontline providers. The majority of health facility directors came from district (69.0%) and public hospitals (65.6%).

**Table 1.**
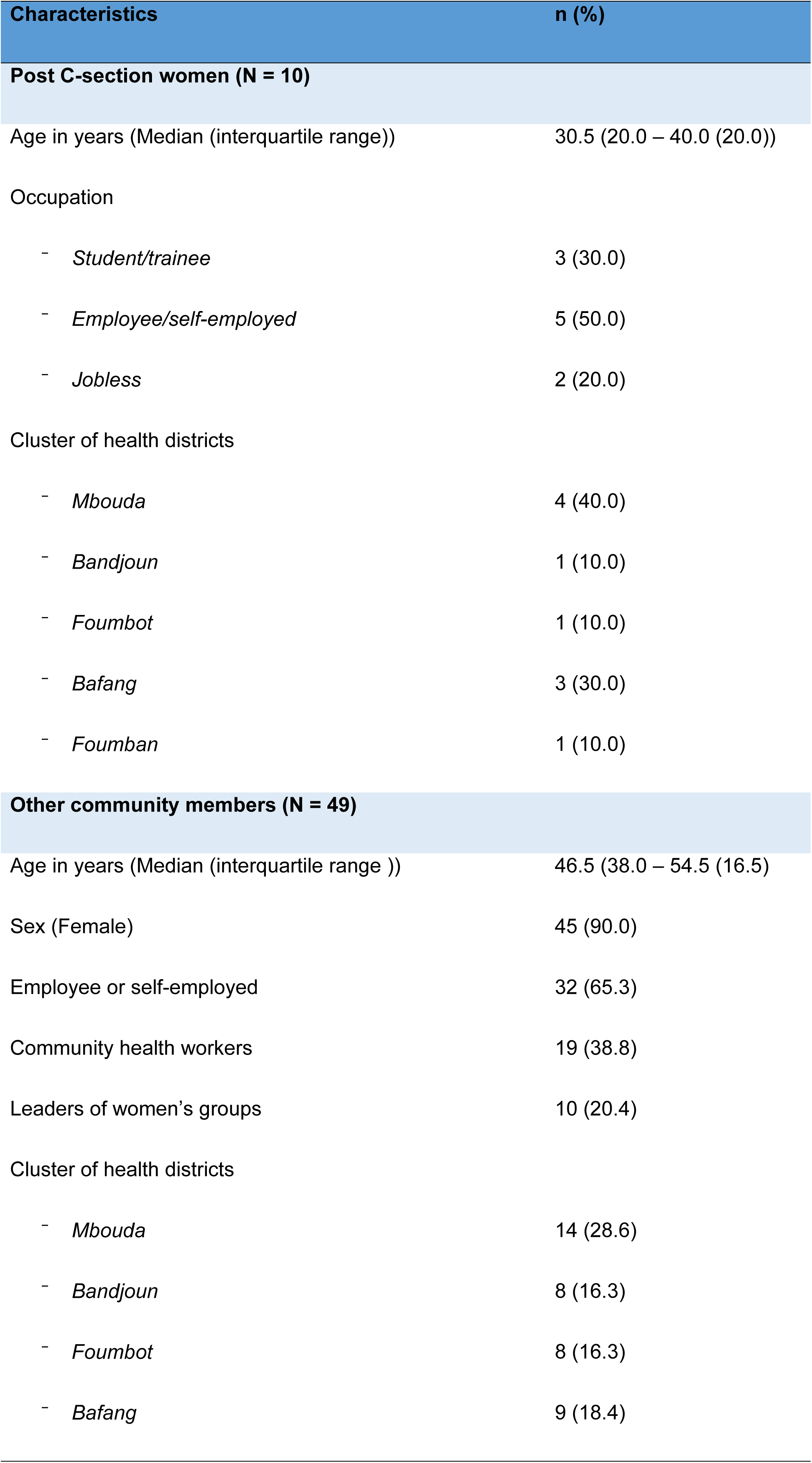
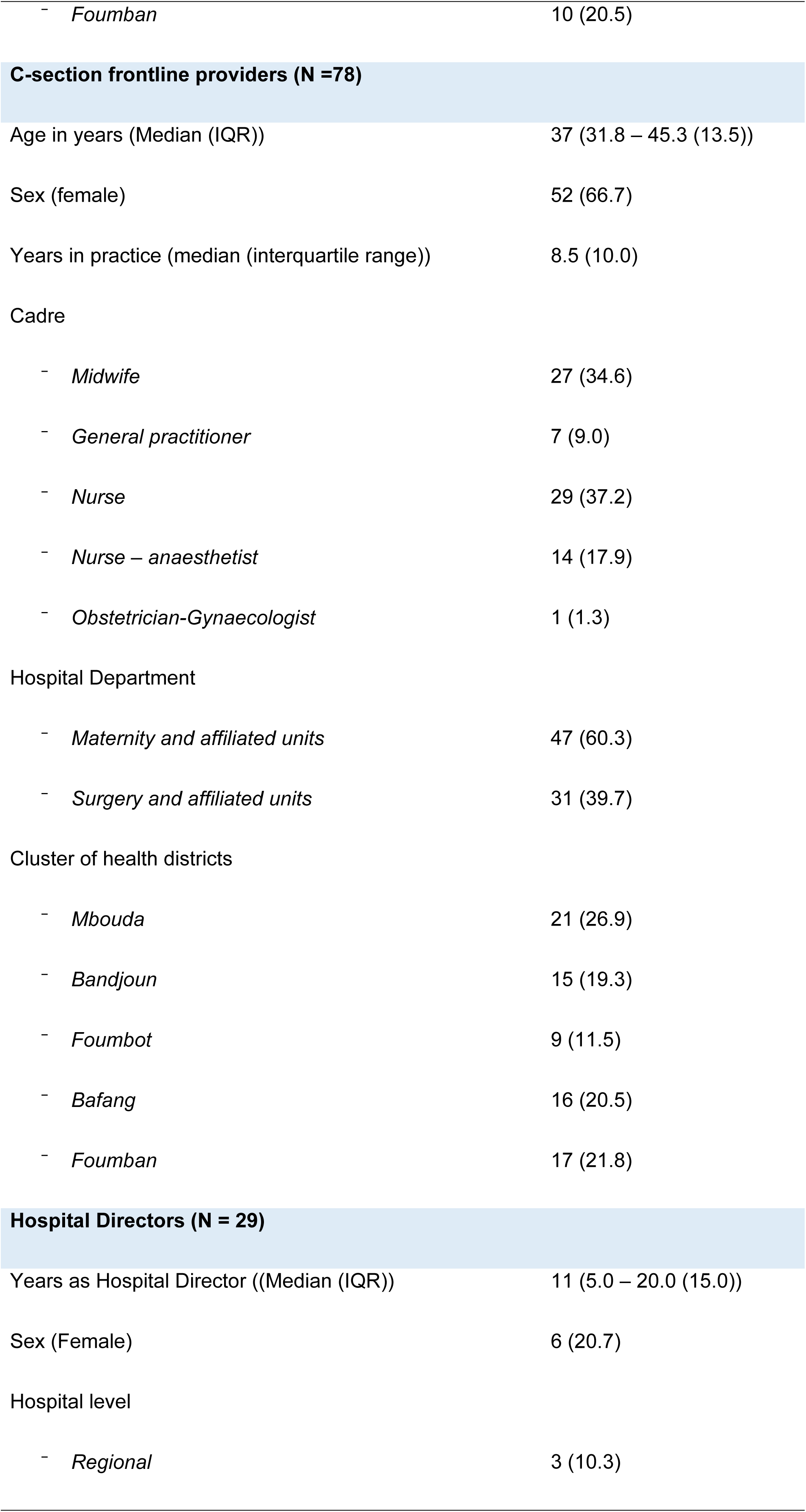
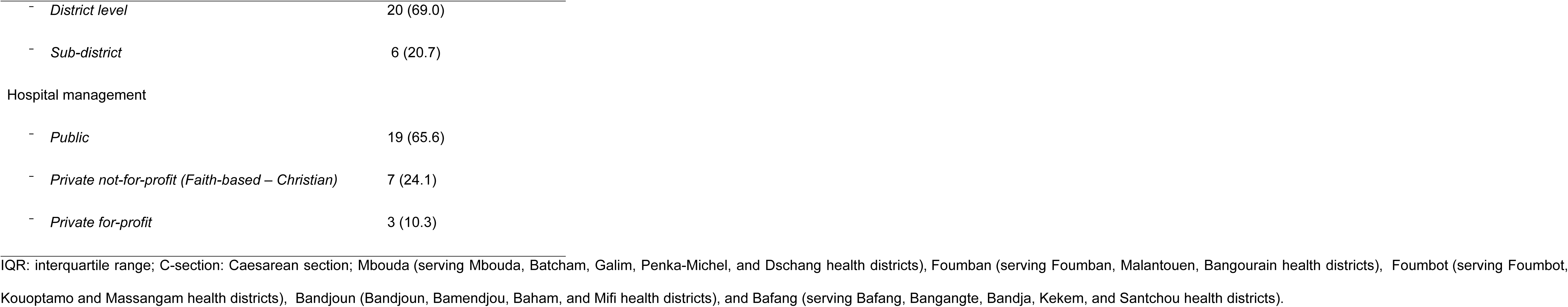
Characteristics of participants (N = 166).

| Characteristics | n (%) |
| --- | --- |
| Post C-section women (N = 10) |  |
| Age in years (Median (interquartile range)) | 30.5 (20.0 – 40.0 (20.0)) |
| Occupation |  |
| - Student/trainee | 3 (30.0) |
| - Employee/self-employed | 5 (50.0) |
| - Jobless | 2 (20.0) |
| Cluster of health districts |  |
| - Mbouda | 4 (40.0) |
| - Bandjoun | 1 (10.0) |
| - Foumbot | 1 (10.0) |
| - Bafang | 3 (30.0) |
| - Foumban | 1 (10.0) |
| Other community members (N = 49) |  |
| Age in years (Median (interquartile range )) | 46.5 (38.0 – 54.5 (16.5)) |
| Sex (Female) | 45 (90.0) |
| Employee or self-employed | 32 (65.3) |
| Community health workers | 19 (38.8) |
| Leaders of women’s groups | 10 (20.4) |
| Cluster of health districts |  |
| - Mbouda | 14 (28.6) |
| - Bandjoun | 8 (16.3) |
| - Foumbot | 8 (16.3) |
| - Bafang | 9 (18.4) |
| - *Foumban* | 10 (20.5) |
| **C-section frontline providers (N =78)** |  |
| Age in years (Median (IQR)) | 37 (31.8 – 45.3 (13.5)) |
| Sex (female) | 52 (66.7) |
| Years in practice (median (interquartile range)) | 8.5 (10.0) |
| Cadre |  |
| - *Midwife* | 27 (34.6) |
| - *General practitioner* | 7 (9.0) |
| - *Nurse* | 29 (37.2) |
| - *Nurse – anaesthetist* | 14 (17.9) |
| - *Obstetrician-Gynaecologist* | 1 (1.3) |
| Hospital Department |  |
| - *Maternity and affiliated units* | 47 (60.3) |
| - *Surgery and affiliated units* | 31 (39.7) |
| Cluster of health districts |  |
| - *Mbouda* | 21 (26.9) |
| - *Bandjoun* | 15 (19.3) |
| - *Foumbot* | 9 (11.5) |
| - *Bafang* | 16 (20.5) |
| - *Foumban* | 17 (21.8) |
| **Hospital Directors (N = 29)** |  |
| Years as Hospital Director ((Median (IQR)) | 11 (5.0 – 20.0 (15.0)) |
| Sex (Female) | 6 (20.7) |
| Hospital level |  |
| - *Regional* | 3 (10.3) |
| - District level | 20 (69.0) |
| - Sub-district | 6 (20.7) |
| Hospital management |  |
| - Public | 19 (65.6) |
| - Private not-for-profit (Faith-based – Christian) | 7 (24.1) |
| - Private for-profit | 3 (10.3) |

IQR: interquartile range; C-section: Caesarean section; Mbouda (serving Mbouda, Batcham, Galim, Penka-Michel, and Dschang health districts), Fouban (serving Fouban, Malantouen, Bangourain health districts), Foubot (serving Foubot,
Kouoptamo and Massangam health districts), Bandjoun (Bandjoun, Bamendjou, Baham, and Mifi health districts), and Bafang (serving Bafang, Bangangte, Bandja, Kekem, and Santchou health districts).

### Problem overview

All participants in the co-design process endorsed the explanations of shortcomings and challenges in the routine practice of IC and debriefing for C-section in the West Region, as identified in our formative study (39,40,50). Reported issues with IC included: limited women’s autonomy during interactions, poor content, domination of provider’s motivation by protection against lawsuits at the expense of women-centred care, and unconducive overall hospital professional context. The main issue with debriefing was its systematic omission. During discussions, they highlighted additional site-specific issues and underscored key aspects of the verbal reports of the formative assessment. One notable topic was hospital governance, described as unsupportive by c-section providers while women and community members stressed that low and irregular wages, coupled with tolerance of malpractice, were the primary drivers of poor consent and debriefing practices. Providers framed the human resource gap as both understaffing and skill-related deficit, while women and community members specifically highlighted their concern regarding the practice of allowing trainees to deliver unsupervised C-section care. On financing, all the groups of stakeholders identified the expensiveness of C-section as a major cause of reluctance toward the procedure; They further called for the immediate inclusion of c-section in the universal health coverage benefits package. Indeed, given that almost all clinical services are paid for from out-of-pocket money, the lack of cash to pay before or immediately after the c-section commonly led to strong rejection of the procedure regardless of the indication.

C-section providers acknowledged that the delivery of IC and debriefing was largely absent in most public hospitals, and where present, it was often marked by negligence (refusal to answer women’s questions), corruption (contingent upon bribery), lack of empathy (providers being harsh on women in pains and distress), and disrespectful treatment (offensive language). They further underscored the need for robust IC and debriefing practices to protect them and the hospital from litigation. Both providers and community groups emphasized the importance of giving women written discharge advice after C-section. Across the 20 health districts, women and community members consistently stressed the need to provide psycho-emotional support to women undergoing C-section, given that the procedure is deeply at odds with prevailing reproductive norms and is frequently experienced as a personal failure (50).

Participants in both groups suggested practices that could facilitate women’s autonomous decision making for IC and strengthen their control towards personal health information during postoperative debriefing, within a context shaped by patriarchal social norms and financial guardianship of husbands prevailing in the region. We noted group-specific suggestions with C-section providers and older women favouring compulsory validation by the husband or a male parent while younger women upheld that IC and debriefing were the woman’s exclusive prerogative. The agreed middle-ground approach was a blended and flexible format whereby the woman chooses her most suitable option, and the relative she designates is counselled to support her decision and to co-sign the IC form as a witness.

### Pre-workshop motivations, perceptions, and experiences of participative collaboration Stakeholders insights on the co-design using the PROSECO framework

#### Dimension 1: Participation

The attendance was optimal, exceeding 95% across all stakeholder groups. Notably, there was no attrition during the deliberative process. A small number of clinical providers were briefly called out to address emergencies in their respective wards. The three categories of stakeholders actively participated in the codesign process adopting the methods used with enthusiasm.

Regarding partnership, pre-existing ties among participants within each workshop group enhanced interactions. As clusters were delineated along geospatial and socio-cultural lines, a majority of clinical providers and community members shared long-standing commonalities. This familiarity often led to spontaneous shifts to local dialects during sessions. Among C-section providers, involvement was underpinned by a collective professional identity and a shared grievance regarding the unconducive hospital environments to standardized consent and debriefing protocols. This group further coalesced around a rejection of traditional rites performed by women and families just before emergency C-sections. Furthermore, they collaborated to advocate for medicolegal protection against unsubstantiated malpractice litigation. For women and community members, the most common cross-cutting goal was to contribute to improving the quality of C-section care in public hospitals. The hospital directors, in turn, closely examine the contextual relevance of the intervention within the existing regulatory framework. For instance, they focussed in-depth on the medico-legal underpinnings of the guidelines for obtaining IC for emergency life-saving C-sections for referred women.

Both community stakeholders and C-section providers reported being highly motivated to contribute to the co-design. The former expressed a profound need to amplify their experiential perspectives and ensure that their nuanced care preferences were integrated into high-level policy dialogues (Figure 1). Among the latter group, C-section providers from public hospitals highlighted how they learned from their counterparts from private faith-based hospitals who willingly shared their practices regarding consent and debriefing.

Hospitals Directors deplored the absence of municipal authorities, religious authorities and traditional healers. In their opinion, municipal authorities who are the statutory chairs of the management committee of public hospitals would have eased the required policy change and additional resource allocation if they had been involved. Likewise, senior religious authorities would have done the same for faith-based hospitals. Regarding traditional rulers who were blamed by C-section providers for delaying IC for emergency procedures could have seized the opportunity to be listened to and engage in a constructive partnership with clinicians.

#### Dimension 2: Experiential

The research funding permitted the procurement of necessary materials and facilitated the uninterrupted execution of the co-design modules. Community stakeholders expressed deep satisfaction regarding the “democratisation” of the design of clinical services manifested by the integration of grassroots perspectives into maternity care protocols. Stakeholders from all groups demonstrated a good understanding of their roles in the co-design with respect to anticipated outcomes. The intervention was perceived to have high contextual relevance, as it directly addressed clinical concerns inherent to routine professional practice or the expectations and lived experiences of healthcare consumers. Likewise, the contextual acceptability and perceived clinical utility of the intervention were high, as evidenced by the spontaneous immediate buy-in by some clinicians at the conclusion of the workshops in view of implementing in their hospitals. Within each cluster, numerous C-section providers proactively sought copies of the co-designed IC instruments and debriefing checklists for immediate integration into their routine clinical workflows.

Regarding process efficiency, a majority of participants noted that sessions were characterized by delayed start and frequently exceeded allotted duration. These deviations were a deliberate methodological choice to accommodate participants traveling from distant geographic locations and to uphold deliberative inclusivity. Restricting the duration of these sessions would have risked the marginalisation of minority voices and compromised the exhaustive exploration of stakeholders’ insights. The pedagogical introduction, the synthesis of formative research findings, and the facilitation were highly appreciated by participants. Furthermore, the diverse and transparent nature of the deliberations were highly regarded. Stakeholders reported enjoying the climate of mutual respect, characterized by the equitable consideration of divergent opinions, the scrupulous validation of shared lived experiences and unrestricted involvement in the deliberative process.

#### Dimension 3: Context

With respect to feasibility, stakeholders perceived that the minimal financial requirements of the intervention would significantly increase its feasibility and the likelihood of successful scale-up. A consensus emerged that the co-designed instruments possessed high contextual and cultural salience. Furthermore, participants identified formal endorsement by regional public health authorities as a pivotal requirements for successful and sustainable implementation. This co-design process was bolstered by several critical contextual determinants. Firstly, it benefitted from the prevailing sociopolitical momentum favouring participatory, rights-based frameworks in public governance, coupled with an escalating public discourse surrounding alleged surgical malpractice in emergency obstetric care. Secondly, the intervention was situated within the interface between modern and traditional medical paradigms characteristic of the region. Finally, the high fertility rate in the Region renders C-section a high-stakes procedure; communal perceptions often associating the procedure with a diminished reproductive trajectory, further intensifying the need for culturally sensitive clinical protocols. On the contrary, hospital Directors indicated that the usually silenced unfair competition and blame shifting culture between hospitals was not discussed during the intervention crafting phase though it negatively impact IC and debriefing. To them, high professional standards regarding competition between hospitals, units and providers were key prerequisites for genuinely context-friendly co-design,

#### Dimension 4: Impact

The co-design process integrated evidence from the formative research with stakeholders experiential knowledge. Stakeholders involved in a process of collaborative sense-making, synthesizing these empirical data with their professional expertise or experiential knowledge to craft intervention components. The way in which the co-designed intervention would be integrated and maintained in the health system is depicted in figure 3. The anticipated mechanisms of change of the interventions components were deemed straightforward by participants and are presented in Table 2. The hospitals directors believed the reach of the intervention could easily encompass the whole West Region of Cameroon. Nevertheless, they underscored the threat represented by the prevailing cross-cutting poor quality of clinical care and services that would hinder the impact of an intervention focusing solely on C-section.

**Figure 3.**
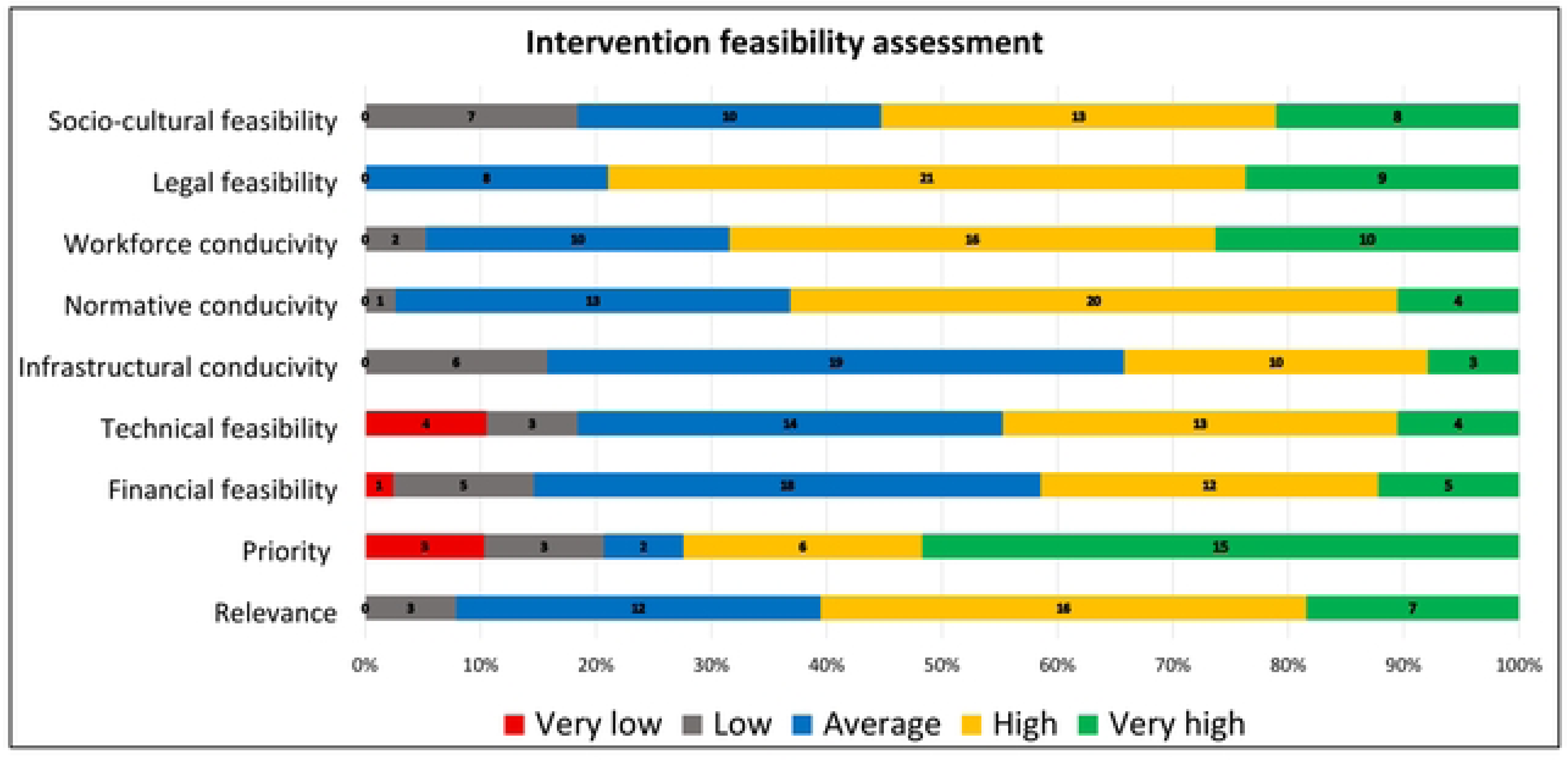
Hospital directors’ assessment of the feasibility of a co-designed hospital-based intervention to improve the quality of informed consent and debriefing in West Cameroon 2026. N = 29. *Ratings: 1 = very low; 2 = Low; 3 = fair; 4 = high; 5 = Very high*

**Table 2.**
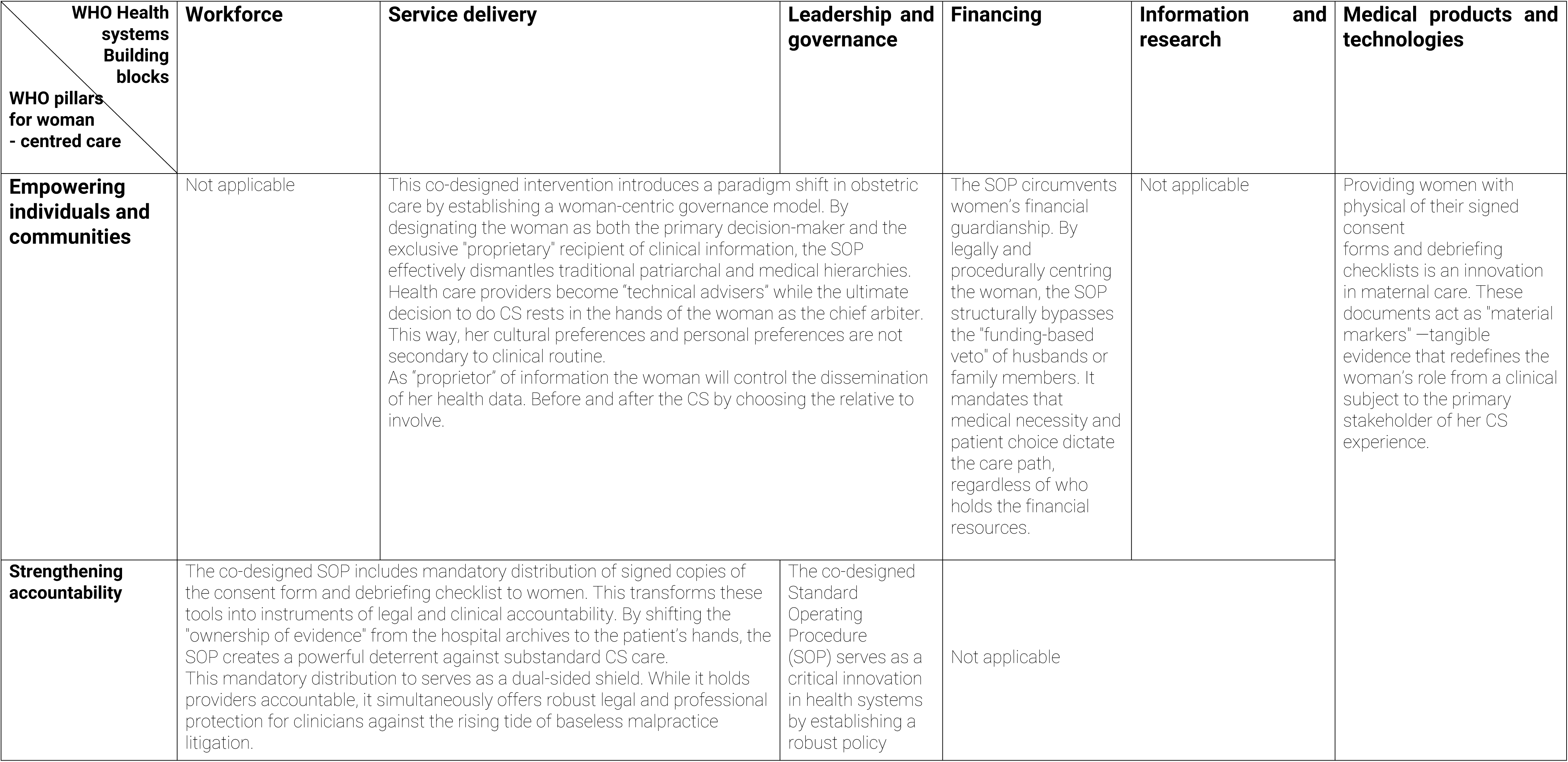
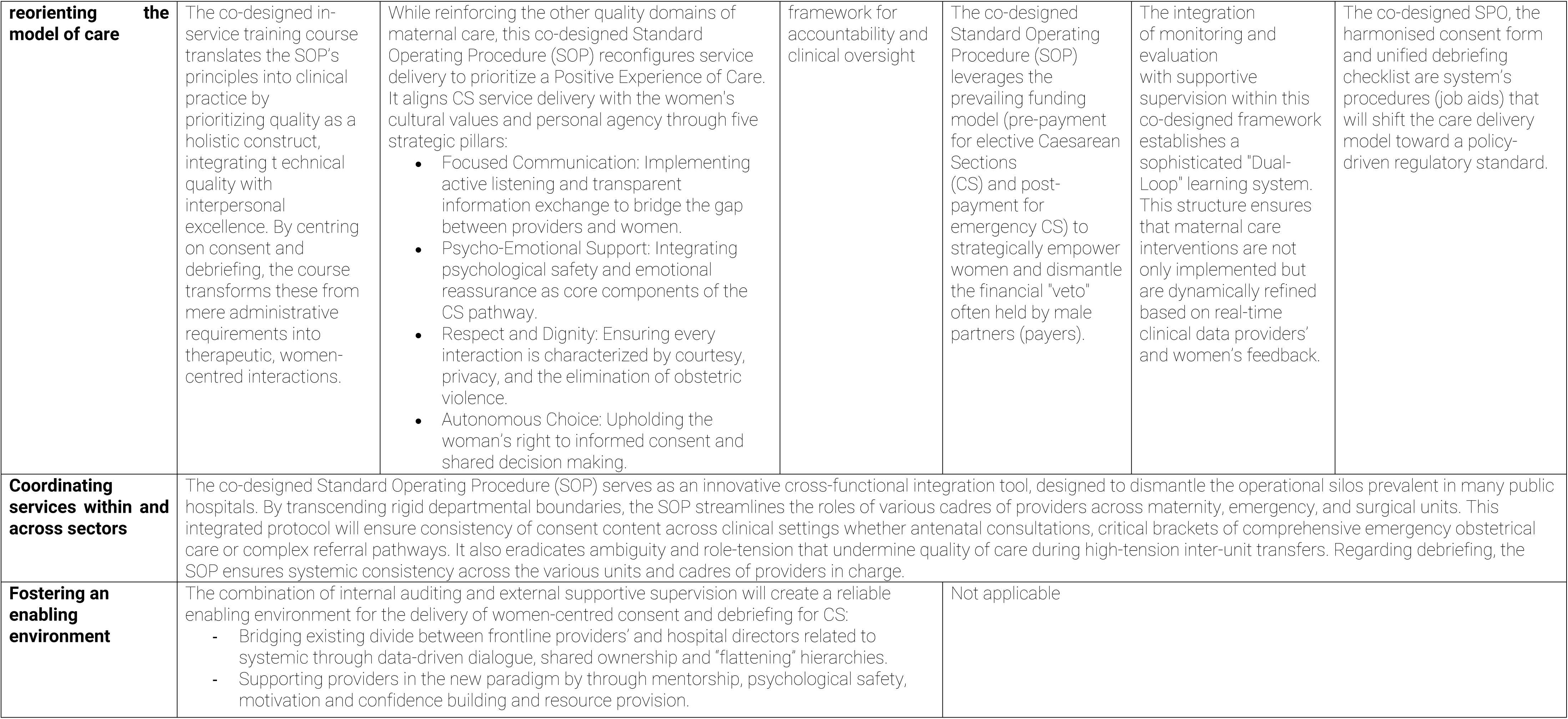
Intersections between co-designed interventions, WHO health systems buildings blocks, and WHO strategic pillars for people-centred care.

#### Dimensions 5: Delivery

The major outputs of this co-design process are summarized using the TIDieR template (Table 3) and supportive instruments (the standard operating procedure, the consent form and debriefing checklist) are available as supplementary files (Supplement 3).

**Table 3.** TIDieR description of intervention components.

|  |  |
| --- | --- |
| **Name** | **Introduction of a standard informed consent form and a unified post-operative debriefing checklist for caesarean section in the West Region of Cameroon.** |
| **Goal** | Improve the quality of informed consent and debriefing for caesarean section through a more woman-centred communication approach |

| Components | In-service training | Adoption of SOPs, consent form & debriefing checklist | Internal auditing | Supportive supervision, monitoring & evaluation |
| --- | --- | --- | --- | --- |
| **Materials** | Meeting room<br>Teaching slide decks and handouts<br>SOPs; Consent form, Debriefing checklist | Policy/guidelines document | Regular (every 14 days) report form<br>Communication allowance | Intervention roadmap<br>Transport, per diems and communication and printing fees<br>Report forms |
| **Methodology** | Single regional all-day induction seminar<br>In-facility one-day training | Managerial/executive decision-making | Involvement, motivation & support of C-section providers<br>Regular checks of completion rates of consent forms and Debriefing checklists<br>Regular phone checks with post-c-section women (& Accompaniers) after discharge | Monthly supportive supervisions (field trips)<br>Bi-weekly field reports<br>Mixed research methods |
| **Person in charge** | - Head of the Regional Sexual and Reproductive Health Unit (Regional Delegation for Public Health) | - Hospital Directors<br>- Regional Delegate for Public Health | - Hospital champions | - Monitoring officer (Head of the Regional Sexual and Reproductive Health Unit (Regional Delegation for Public Health)) |
| **Activities** | - Logistics for the seminars<br>- Planning & preparation of teaching<br>- All-day adult learning activities | Formal administrative decision | Check-in meetings with frontline practitioners<br>Report write up<br>Phone calls with post-caesarean section women after discharge | Progress review<br>Records review<br>Quantitative assessment using ad hoc indicators<br>In-depth interviews with stakeholders<br>Report to be shared with the MoH other Regions<br>Manuscript and policy brief write up |
| **Place** | Regional Delegation headquarters | Hospitals | Hospitals | Hospitals and Regional Delegation headquarters |
| **Start date** | A soon as possible | Immediately after the Regional induction seminar | The week following the in-facility training | From the selection of facility champions (Monitoring)<br>At the end of 14<sup>th</sup> week (Evaluation) |
| Frequency and duration | Once | Once | Every 10 days for 12 consecutive weeks | Bi-weekly for 12 Weeks (Monitoring)<br>Once (Evaluation) |
| Adaptation to context | As and when needed | As and when needed | As and when needed | As and when needed |

SOP: Standard operating procedure; MoH: Ministry of Health.

### TIDieR description of the intervention, stakeholders’ contributions and supporting materials

Table 3 summarises the four components of the co-designed intervention using the TIDieR framework, which are: (1) an in-service training module, (2) a standard operating procedure using a harmonised consent form and a unified debriefing discharge checklist; (3) an internal auditing scheme, and (4) an external supervision monitoring and evaluation scheme. The harmonised co-designed IC form, the unified debriefing discharge form and the related standard operating procedure are described as a supplementary material (supplement 3).

#### Component 1: In-service training module

This training module consists of a whole-day on-site seminar using adult-teaching approaches on the following topics: disrespect and abuse in maternity care; medico-legal requirements of c-section; respectful maternity care, women’s sexual and reproductive rights, and psycho-emotional support in maternity care. The target trainees are clinicians directly involved in C-section care (antenatal, labour ward, emergency, surgical theatre, and post-natal units). The sexual and reproductive health department of the Regional Delegation of Public Health was identified to supervise the training using validated teaching materials and involving external consultants as needed. Given that the co-designers were not licensed teachers, they were not expected to design the teaching module like experts. They outlined the goals of the teaching, expected results, sites, duration and more specific pedagogic aspects were left to the regional delegation and its academic partners

Regarding the medico-legal aspects, C-section providers highlighted a strong need for capacity building to navigate the rise in litigations in maternity care in Cameroon. Concerning psycho-emotional support, women and community members deplored the lack of compassion and psycho-emotional support to address the major distress women experience before, during and after C-section, and strongly suggested the recruitment of clinical psychologists in every maternity. Concurrently, C-section providers and hospitals directors unanimously pointed to the absence of psychologists from hospitals and the limited proficiency of maternity staff in delivering psycho-emotional care. In the face of normalised disrespectful practices in c-section and maternity care at large, women and community members emphasized the utmost necessity to make providers knowledgeable and respectful of women’s sexual and reproductive rights despite the asymmetric power dynamics in clinical settings.

#### Component 2: Standard operating procedure using a harmonised consent form and a unified debriefing discharge checklist

Mindful of routine practices in maternity and surgical theatres and factoring in women’s main preferences, the standard operating procedure to seek and document IC and carry out debriefing was framed as a cascade specifying the actions and actors of each step in both emergency and elective settings. On the woman’s side a third party of her choice has to be involved as a witness and contact person to ease the continuity of care. This “co-consenter” is not entitled to override the woman’s decision. The IC form and debriefing discharge checklist were attached to the standard operating procedure document. Women and other community participants emphasised the psycho-emotional support to be offered by provider’s during consent for emergency C-section and debriefing. Besides, they wanted providers to ensure women have unrestricted access to their chosen alternative medical practitioners including herbalists, traditional healers and religious leaders.

On the grounds of the limited administrative support for consistent practice of IC and debriefing in public health facilities, c-section providers pushed hard for adoption of clinical guidelines to streamline practices. The idea of a standardized operating procedure using an IC form and a unified debriefing checklist was suggested by hospital managers to ensure consistency across the various cadres of C-section providers in charge of consent and debriefing in the Region. On their side, women and other community members insisted on the necessity to receive a detailed discharge notice as a reminder of key post-operative medical instructions/advice given the barriers post C-section women face to access post-natal visits. In addition, it is anticipated that the document will help women securing husbands’/male partner’s support in observing post c-section recovery restrictions at home.

#### Component 3: Internal audit

It was agreed that every hospital offering C-section would designate a champion in charge of: (1) involving, motivating and supporting C-section providers in the provision of consent and debriefing; (2) carrying out regular checks of completion rates of consent forms and debriefing checklists; (3) liaising through phone checks with post C-section women (and accompaniers) after discharge. Their designation should consider their leadership position in the hospital, their demonstrated motivation towards respectful maternity care and their voluntariness to assume the role. The champion would organize regular check-in meetings with C-section providers and write up a monthly progress and analytical (SWOT) report on the state of delivery of consent and debriefing alongside suggestions to address shortcomings to the attention of the hospital director. This designation of a champion was suggested by senior C-section providers who presented it as the most effective way to root the intervention in routine practice. They explained that most successful public health programmes and units implemented in hospitals owe their success to the performance of dedicated champions locally referred to as “focal person”.

#### Component 4: external supervision, monitoring and evaluation

To ensure (1) effective coordination and oversight of these “new” services across the Region, (2) achieve their integration in the routine package of sexual, reproductive, maternal and adolescent health services and (3) deliver on accountability to communities, it was decided that the Regional Delegation for Public Health would carry out monitoring and evaluation of the intervention in hospitals alongside monthly on-site supportive supervision. Besides routine monitoring the tasks (records and progress review), the Delegation will engage community leaders, hospital champions and directors about the issues identified by the internal audits. Post C-section women and communities representatives strongly supported this external oversight which they viewed as an accountability mechanism to address disrespect, abuse and neglect before and after C-section. C-section providers further stated that this external supervision by the top public health authorities in the Region would pressure hospital directors towards an enabling environment.

### Intersection between the intervention’s components and the WHO health system buildings blocks and strategic pillars for integrated people-centred services

Intersections between co-designed interventions, WHO health systems buildings blocks, and WHO strategic pillars for people-centred care are depicted in supplement 4:

### Scalability of the interventions

#### Feasibility assessment by hospital directors

The applicability of the co-designed intervention in the various hospital contexts has been examined by hospital directors, and their perceived ratings on a five-point Likert’s scale (1 = very low; 2 = Low; 3 = fair; 4 = high; 5 = Very high) are summarized in Figure 2. The median (Interquartile range) tentative duration to implement the intervention estimated by 23 hospital directors was 3 (3) months. The scalability of this multicomponent intervention in the West Region of Cameroon hinges not only on its institutional adoption but also on its applicability.

#### Facilitators and barriers to implementation of the co-designed intervention

As prospective leaders of the implementation of the co-designed intervention, hospital directors provided critical insights on the implementation landscape by identifying facilitators and barriers. In supplement 5, we summarized the major facilitators and barriers they put forward in a cross-reading with the WHO strategic pillars for people-centred and integrated care services.

## Discussion

### Main outputs

We reported on a co-design process that led to the development of a hospital-based multicomponent intervention to improve IC and post-operative debriefing for C-section in the West Region of Cameroon. This process was informed by earlier formative research on routine practices in the region and co-developed by more than 166 stakeholders, including women who had undergone C-section, other community members, frontline clinicians, and hospital leaders. The resulting intervention, presented in the TIDieR format, is made up of four components: an in-service training curriculum, a harmonized consent form and unified debriefing checklist, a standard operating procedure, and a supportive supervision scheme for health care providers. The in-service training component is a one-off workshop delivered to C-section frontline providers on rights-based respectful maternity care (including C-section) and a hands-on introduction to the provision of consent and debriefing following the co-designed standard operating procedure. The harmonised consent form and the unified debriefing checklist are a blend of universal requirements and context-friendly cultural adaptations framed in a user-friendly format. The supportive supervision scheme is centred on a hospital champion promoting and monitoring the use of the co-designed tools with the support of the hospital director to address eventual issues and the regional sexual and reproductive health supervisory team for evaluation. the co-design approach was chosen to ensure that the intervention is intrinsically aligned with both the regional sociocultural norms and the operational exigencies of the hospital environment. Feedback from participants on the co-design indicated that core requirements for meaningful participation were efficacious, and the intervention was perceived as feasible by intended lead implementers.

### Findings in context

#### Process

Our co-design process adhered to the recommended four-stage cascade (preparation/planning, design/conducting, evaluation/reporting and implementation) for the co-creation of public health interventions (33,51). In 2025, Vargas et al. published a systematic review on co-design processes, models and frameworks in public health in which they reported that over half of articles utilised the same sequence (32). The co-design process described here also accounts for the local socio-cultural norms, the regulatory framework and hospital systemic environments, thereby meeting key elements for successful implementation of public health intervention in low- and middle-income countries (52).

#### Participants and context-friendliness

The number of community members involved in our co-design process places it among the relatively small number of studies reporting more than 100 participants (53). This large number was required to account for the multiple professional perspectives and the diverse sociocultural contexts in the region. The spectrum of participation and contribution of non-academic stakeholders in action research spans from non-participation to control of research. Our study met the middle ground by ensuring participants crafted and approved interventions (33). This ensured consideration of all aspects of the interactions between women and C-section providers during consent and debriefing (54). While most of the health guidelines and normative products from community engagement articles featured in the systematic review by Bohren et al. in 2025 had a national or supra-national scope, our intervention primarily targets one region (53). This “narrow” scope was dictated by the pronounced context-specific nature of our co-design process which needed to account for the diverse cultural norms. Although local specificities exist, the West Region belongs to one of the four overarching ethno-cultural zones of Cameroon. We believe that despite that this version of the intervention could not fit the whole country as a one size-fits-all solution, the methodology we used could be replicated in the three other ethno-cultural zones to develop versions relevant to their contexts(55,56). As the majority of action research projects, our process involved the people affected by the intervention and providers to develop a service improvement intervention (32,53). This approach aligns with the concept of localisation which is gaining momentum on global, regional and national stages for effective policy making (57). Furthermore, our co-designed intervention meets a key implementation pre-requisite highlighted by the literature as it is in line with Cameroon’s national public health strategy aimed at upgrading quality and coverage of clinical services (58).

#### Intervention components

Multi-component interventions based on standardised protocols, IC checklists, providers’ training and visual (paper or multimedia) aids have been shown to improve recollection of IC and overall caesarean birth experience (59–61). In 2020, in a before-and-after study, Zethof et al. reported that an intervention in a rural mission hospital in Malawi combining the introduction of an IC checklist, a training of c-section providers on communication and a wall poster reminder significantly decreased the incompleteness of IC and improved women’s recollection of common c-section complications (59). In comparison, our co-designed intervention comprises a training module on communication with women, and a checklist but not the wall poster reminder. We hypothesise that he reminder role played by the latter in Zethof et al.’s work would be replaced in our co-designed intervention by the constant engagement of c-section providers by the designated hospital champion.

Single-component interventions based on communication aids or training have also been reported as efficacious in improving consent for c-section. In 2020, Truong et al. in randomised controlled trial in Melbourne found no significant difference between the use of an adjunct written pamphlet and an adjunct multimedia information during consent in improving women’s knowledge for elective c-section(62). Nevertheless, both tools significantly increased women’s knowledge between the IC day and the day of the surgery two to four weeks later. It is not surprising that our co-designers did not suggest written information to support IC not only because 80% of c-section in the Region were emergency procedures and that they reported no understanding issues with their routine practice of IC for elective procedures. Likewise, no multimedia intervention was suggested during our co-design because of its absence from the provider-patient interaction in the Region and the numerous challenges making its adoption unrealistic (63) .Interventions based on training alone reported as successful in improving IC and debriefing(64). The in-service training component of our intervention shares key characteristics with most of the training packages featured by Yargawa et al. in their scoping review published in 2024 on the content and design of respectful maternity care trainings in sub-Sahara Africa (64). The most common training topics identified in their review were similar to those included in the training we co-designed: (i) effective communication, (ii) prospective provision of information and seeking informed consent; (iii) ensuring continuous access to family and community support (iv) patients’ rights and (iv) patient-centeredness. Likewise, the most common training format identified in the review was workshop-based, in-service and targeting frontline women care providers, similar to our intervention. Furthermore, the most common delivery method of the trainings in their review was a one-off workshop. Last, 90% of the reviewed studies assessed the impact of training as our intervention proposes to do. Overall, our training aligns with approaches and practices reported elsewhere in sub-Saharan Africa. Notably, none of the 27 studies in Yargawa’s review were conducted in Cameroon (64).

Studies reporting interventions to improve the quality of debriefing in Africa are rare (12,65). In Belfast, United Kingdom; Dougan et al. reported their successful 6-month implementation of a three-component hospital-based intervention to improve women’s “full clarity” on post C-section debriefing (66). Their components were similar to the ones we co-designed: the promotion of debriefing as a significant aspect of care (diverse reminders in routine practice), the introduction of a debrief form and education of junior doctors in methods of debriefing. Over six months, they achieved progress on “full clarity” from 20% to 60%. Nevertheless, they acknowledged that their late engagement of senior managerial team members and the whole healthcare team including midwives caused several implementation problems including delays. Our co-designed intervention has been approved by hospitals directors to avoid such issues.

### Policy and global implications

This research helps with the improvement of respectful maternal care, an area where we still have too few interventions. Moreover, the knowledge developed through this deliberative process constitutes a resource that Cameroonian authorities may leverage for the broader health system transformation agenda, such as the hospital reform (under the umbrella of the 2020-30 health sector strategy) which proposes the implementation of standard clinical protocols and dignified care (67,68). On the global stage, this research addresses a significant knowledge gap by providing empirical evidence for maternal health action research in low- and middle-income countries, as pursued by the World Health Organization’s Integrated People-Centred Health Services framework.

### Strengths and limitations

This co-design helped identify practical ways to improve communication, IC and debriefing practices. Most actors (researchers and participants) were indigenous to the Region, and the concepts and procedures were informed by local knowledge, including lived experience, sociocultural norms, and prevailing paradigms of female empowerment. Deliberations also ensured that the final outputs accounted for local clinical and hospital institutional environments. To our knowledge, this co-design initiative is the first in the West Region of Cameroon to yield a ready-to-implement intervention aligned with the global movement towards respectful, woman-centred and integrated reproductive health care. It further innovates in describing a methodology to contextualise the principles of IC and debriefing in a non-western collectivistic African community.

Nevertheless, our co-design process has four main limitations. First, the evaluation of the co-design process was limited to using the PROSECO criterion-based framework to synthetise participant’s opinions on its overall implementation. There was no external process evaluation conducted. Second, the implementation of the intervention has not been formally tested, meaning its potential effectiveness in changing consent and debriefing practices is unknown. Third, the intervention has no community-based component to address demand-side issues that negatively impact IC and debriefing practices, such as delays in reaching health care facilities. Fourth, the validation of the final version of the intervention did not involve the socio-cultural gate keepers as traditional rulers, religious authorities and political opinion leaders.

### Future research

Future research could focus on piloting the intervention in the region to evaluate its clinical effectiveness and assess its potential for scalability. Besides, research using the approach we reported could be conducted in other settings.

### Conclusion

This participatory co-design process yielded a context-specific, multi-component intervention that was well accepted and deemed feasible across diverse clinical and community stakeholders. It provides the methodological basis for a practical approach to strengthening of IC and debriefing as core elements of women-centred, accountable maternity care, and warrants implementation.

### Author contributions

|  | JTF | MS | LTD | MM | WCDK | BK | LB | VF |
| --- | --- | --- | --- | --- | --- | --- | --- | --- |
| Conceptualisation | √ | √ | √ | √ |  |  | √ | √ |
| Data collection | √ |  |  |  | √ |  |  |  |
| Data analysis | √ | √ | √ |  | √ |  |  | √ |
| Funding acquisition | √ | √ | √ |  |  |  | √ | √ |
| Project administration | √ | √ | √ |  |  |  |  | √ |
| Writing_original draft | √ |  |  |  |  |  |  |  |
| Writing_review and editing |  | √ | √ | √ |  | √ | √ | √ |
| Approval of the last version | √ | √ | √ | √ | √ | √ | √ | √ |

## Data Availability

No data was generated by this study.

## Supporting information captions

**Supplement 1**. Map of the West Region of Cameroon.

**Supplement 2.** Participants’ pre- and post-workshops opinion collection forms. Co-design of complex hospital-based intervention to improve the quality of informed consent and debriefing in West Cameroon. 2026.

**Supplement 3.** Co-designed standard operating procedures, harmonised informed consent form and unified postoperative debriefing checklist for caesarean section in West Cameroon. 2026.

**Supplement 4.** Intersections between WHO health systems buildings blocks, WHO strategic pillars for people-centred care, and co-designed standard operating procedures for informed consent form and unified postoperative debriefing checklist for caesarean section in West Cameroon. 2026.

**Supplement 5.** Perceived facilitators and barriers to implementation of a co-designed complex hospital-based intervention to improve the quality of informed consent and debriefing in West Cameroon. 2026. N = 29 hospitals directors.

## Notes

### Competing Interest Statement

The authors have declared no competing interest.

### Funding Statement

This work was supported by the Japanese Government through its world innovative and smart education grant, and by the London school of Hygiene and Tropical Medicine through its research degree travel grant.

### Author Declarations

1- The Institutional Review Board of the London School of Hygiene & Tropical Medicine (Ref: 29898) 2- Regional Ethics Committee for Human Health Research of the West Region, Cameroon (Ref: No/984/25/10/2023/CE/CRERSH-OU/VP).

